# The Cancer Burden in the Abu Dhabi Population: A Retrospective Cohort Study

**DOI:** 10.1101/2025.01.18.25320767

**Authors:** Latifa Baynouna AlKetbi, Rawan Ashoor, Ekram Saeed, Reem AlShamsi, Nico Nagelkerke, Noura AlAlawi, Rudina AlKetbi, Hamda Aleissaee, Noura AlShamsi, Ahmed Humaid, Hanan Abdulbaqi, Toqa Fahmawee, Basil AlHashaikeh, Muna AlDobaee, Nayla AlAhbabi, AlYazia AlAzeezi, Fatima Shuaib, Jawaher Alnuaimi, Esraa Mahmoud, Mohammed AlMansoori, Sanaa AlKalbani, Wesayef AlDerie, Amira AlAhmadi, Mohammad Sahyouni, Maha AlAzeezi

## Abstract

**Background:** Cancer is the third-leading cause of death in the United Arab Emirates (UAE). There are few studies on cancer epidemiology in the UAE.

**Methods:** This retrospective study, from 2011 to 2013 until 2023, aimed to estimate the prevalence of a history of cancer and lifetime risk and identify its risk factors.

**Results:** Among 8635 cancer-free participants at the time of screening, 209 were diagnosed with new cancer (2.5%) over the follow-up period, 129 (1.9%) females compared to 80 (0.9%) males. The most common type of cancer was Breast cancer, with an incidence of 18% (37 cases) out of the 209 total cases, all females. The second most common type was Thyroid Cancer, with 31 cases; 24 of them were females, and 7 were males. The third most common type was colon cancer, with 22 cases, 11 females and 11 males. Other types of cancer were found but had lower incidence, such as Adrenal, skin cancer, ovarian, bladder, and gastrointestinal GI cancers. The prevalence was comparable to the latest published report of the Abu Dhabi cancer registry in 2019.

Among the whole cohort, Cox regression analysis showed only age and higher levels of HDL as risk factors for any cancer after screening when including possible risk factors that were assessed at baseline. The risk increases by 4.8% for each year older and 52% for each unit increase in HDL. The area Under the ROC Curve of the developed model for predicting any cancer type is 0.739 (0.707-0.771).

**Conclusion:** This study calls for focused research on different types of cancer to identify significant predictors to aid early diagnosis and management, as well as research on prevention and survival. This study’s results provide critical input to the country’s decision-makers on cancer services and are informative internationally as the risk factors identified are prevalent worldwide and may be targeted with available interventions.

## Introduction

An increasing body of literature describes the many biological processes and pathways, incidence, mortality, and response to treatment for cancer patients[1]. However, more research is needed on geographical and cultural aspects of cancer epidemiology and identifying groups of patients who are different in disease susceptibility as well as treatment response to better target prevention, early detection, and more accurate and personalized therapy[1]

In a systematic analysis of the Global Burden of Disease Study, from 2010 to 2019, global risk factor-attributable cancer deaths increased by 20·4%. Interestingly, the leading risk factors were behavioral, notably smoking, followed by alcohol use and high BMI, which varied by world region and Socio-demographic Index (SDI). In particular, the largest increases between 2010 and 2019 were seen in metabolic risk factors [2].

Cancer is the third-leading cause of death in the United Arab Emirates (UAE) [3], and both smoking and high BMI are highly prevalent. Reducing exposure to modifiable risk factors could decrease cancer mortality and patients’ quality of life worldwide by over 50% [4]. However, estimates for the UAE have yet to be made. The UAE has invested heavily in its healthcare system, including many screening programs for early disease detection and management. This has provided a wealth of longitudinal data for investigating the epidemiology and etiology of different diseases.

This study is part of a large retrospective study in the Abu Dhabi Emirate with a relatively long duration, from 2011 to 2013 until 2023. It aimed to estimate the prevalence of a history of cancer and lifetime risk and possible determinant risk factors.

## Methods

This retrospective cohort study is based on a national health screening program focused on cancer incidence, risk factors, and estimation of its risk in an Arab population of UAE nationals living in the Abu Dhabi Emirate. 8699 cohort participants underwent cardiovascular screening between 2011 and 2013 as part of the national program WEQAYA. This Abu Dhabi emirate-based program targets UAE nationals over 18 to reduce their risks of cardiovascular diseases.

Among all participants, there were 64 who had cancer before the screening date; 37.5% were males and 62.5% were females. Follow-up of the 8635 initially cancer-free individuals was carried out in 2023 by physicians and nurses using an Abu Dhabi-wide health information system, with an average follow-up of 9.2 years (range <1 year-12 years), during which cancer incident cases were identified.

Details of the study’s methodology can be found elsewhere. Variables collected at baseline included demographic data, self-reported health indicators including smoking status, physical activity, preexisting CVD, diabetes, hypertension, and dyslipidemia, and whether participants were taking medication for these conditions. Anthropometric measures included waist and hip circumference, body mass index (BMI in kg/m2), and a single arterial blood pressure reading. . Hematological parameters included non-fasting glucose (mmol/L), total cholesterol, high-density lipoprotein (HDL) cholesterol (mmol/L), glycosylated hemoglobin (HbA1c), vitamin D, and creatinine (12). As some subjects had missing data, only subjects with complete data were included.

### Statistical analysis

Frequencies, descriptive statistics, and Cox survival regression with times of diagnosis of cancer as endpoints. Kapplan-Mier curves were used to study the expression of risk factors related to cancer incidence over time.

## Results

Participants in the national screening program in 2011-2013 had a personal history of cancer of 0.92% in females and 0.55% in males. Over the 12-year follow-up period, the incidence in females was 1.8% and in males 1.9% %. Table 1 shows the total prevalence of cancer over the follow-up years.

**Table 1.**
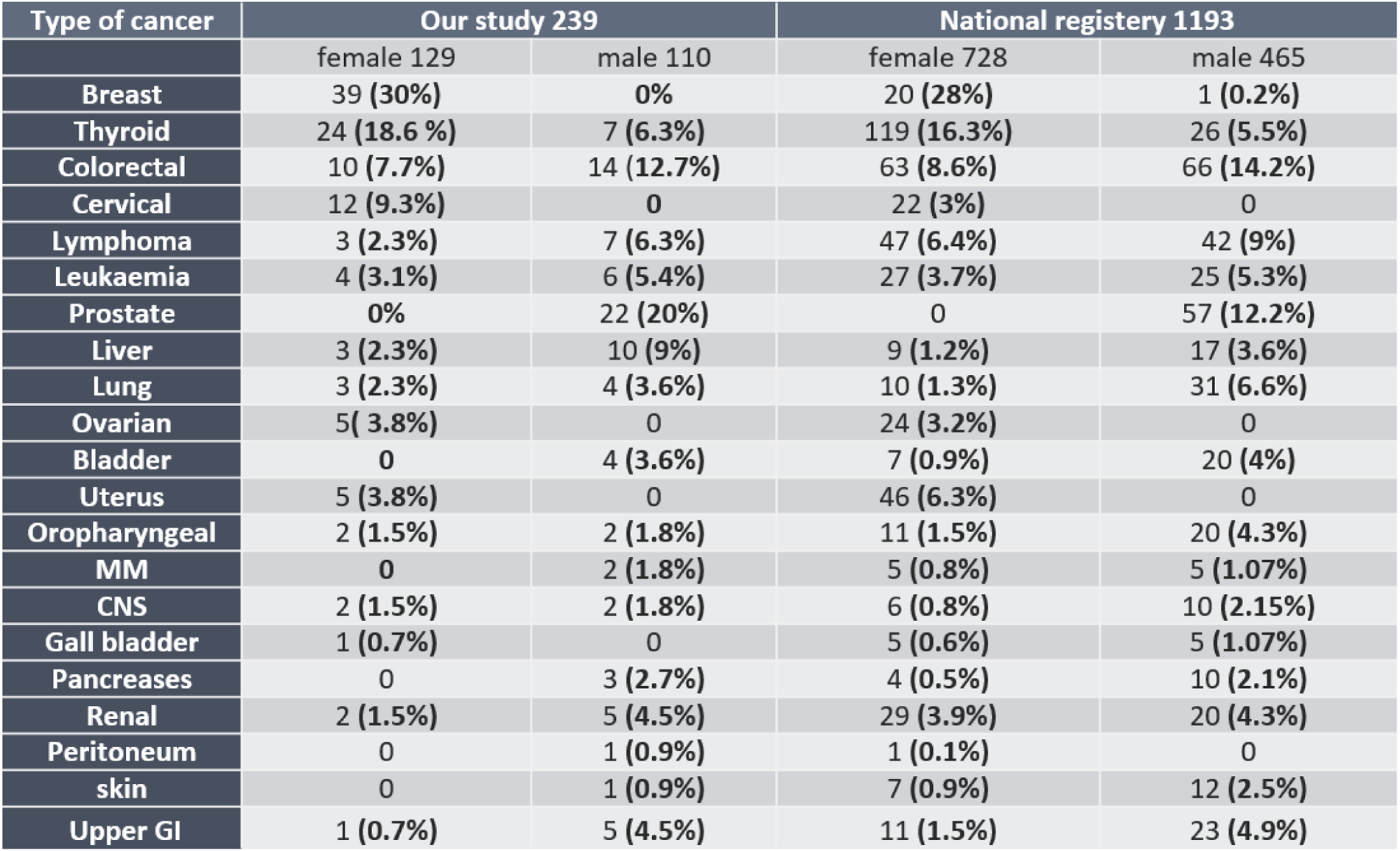

| Type of cancer | Our study 239 |  | National registry 1193 |  |
| --- | --- | --- | --- | --- |
|  | female 129 | male 110 | female 728 | male 465 |
| Breast | 39 (30%) | 0% | 20 (28%) | 1 (0.2%) |
| Thyroid | 24 (18.6 %) | 7 (6.3%) | 119 (16.3%) | 26 (5.5%) |
| Colorectal | 10 (7.7%) | 14 (12.7%) | 63 (8.6%) | 66 (14.2%) |
| Cervical | 12 (9.3%) | 0 | 22 (3%) | 0 |
| Lymphoma | 3 (2.3%) | 7 (6.3%) | 47 (6.4%) | 42 (9%) |
| Leukaemia | 4 (3.1%) | 6 (5.4%) | 27 (3.7%) | 25 (5.3%) |
| Prostate | 0% | 22 (20%) | 0 | 57 (12.2%) |
| Liver | 3 (2.3%) | 10 (9%) | 9 (1.2%) | 17 (3.6%) |
| Lung | 3 (2.3%) | 4 (3.6%) | 10 (1.3%) | 31 (6.6%) |
| Ovarian | 5 (3.8%) | 0 | 24 (3.2%) | 0 |
| Bladder | 0 | 4 (3.6%) | 7 (0.9%) | 20 (4%) |
| Uterus | 5 (3.8%) | 0 | 46 (6.3%) | 0 |
| Oropharyngeal | 2 (1.5%) | 2 (1.8%) | 11 (1.5%) | 20 (4.3%) |
| MM | 0 | 2 (1.8%) | 5 (0.8%) | 5 (1.07%) |
| CNS | 2 (1.5%) | 2 (1.8%) | 6 (0.8%) | 10 (2.15%) |
| Gall bladder | 1 (0.7%) | 0 | 5 (0.6%) | 5 (1.07%) |
| Pancreases | 0 | 3 (2.7%) | 4 (0.5%) | 10 (2.1%) |
| Renal | 2 (1.5%) | 5 (4.5%) | 29 (3.9%) | 20 (4.3%) |
| Peritoneum | 0 | 1 (0.9%) | 1 (0.1%) | 0 |
| skin | 0 | 1 (0.9%) | 7 (0.9%) | 12 (2.5%) |
| Upper GI | 1 (0.7%) | 5 (4.5%) | 11 (1.5%) | 23 (4.9%) |

**Table 2(A).**

| CANCER_BEFORE | Metric | Statistic | Std. Error |
| --- | --- | --- | --- |
| 0 Age at screening | Mean | 40.68 | 0.238 |
|  | 95% Confidence Interval Lower Bound | 40.21 |  |
|  | 95% Confidence Interval Upper Bound | 41.14 |  |
|  | 5% Trimmed Mean | 40.09 |  |
|  | Median | 36.8 |  |
|  | Variance | 245.903 |  |
|  | Std. Deviation | 15.681 |  |
|  | Minimum | 14 |  |
|  | Maximum | 93 |  |
|  | Range | 78 |  |
|  | Interquartile Range | 25 |  |
|  | Skewness | 0.474 | 0.037 |
|  | Kurtosis | -0.832 | 0.074 |
| 1 Age at screening | Mean | 58.9 | 3.056 |
|  | 95% Confidence Interval Lower Bound | 52.58 |  |
|  | 95% Confidence Interval Upper Bound | 65.22 |  |
|  | 5% Trimmed Mean | 59.55 |  |
|  | Median | 62.76 |  |
|  | Variance | 224.146 |  |
|  | Std. Deviation | 14.972 |  |
|  | Minimum | 20 |  |
|  | Maximum | 87 |  |
|  | Range | 67 |  |
|  | Interquartile Range | 15 |  |
|  | Skewness | -0.974 | 0.472 |
|  | Kurtosis | 1.46 | 0.918 |

**Table 2(B).**

| CANCER_BEFORE | Metric | Statistic | Std. Error |
| --- | --- | --- | --- |
| 0 Age at screening | Mean | 37.95 | 0.214 |
|  | 95% Confidence Interval Lower Bound | 37.53 |  |
|  | 95% Confidence Interval Upper Bound | 38.37 |  |
|  | 5% Trimmed Mean | 37.18 |  |
|  | Median | 35.73 |  |
|  | Variance | 196.951 |  |
|  | Std. Deviation | 14.024 |  |
|  | Minimum | 14 |  |
|  | Maximum | 112 |  |
|  | Range | 98 |  |
|  | Interquartile Range | 20 |  |
|  | Skewness | 0.73 | 0.037 |
|  | Kurtosis | 0.137 | 0.075 |
| 1 Age at screening | Mean | 47.45 | 2.43 |
|  | 95% Confidence Interval Lower Bound | 42.54 |  |
|  | 95% Confidence Interval Upper Bound | 52.37 |  |
|  | 5% Trimmed Mean | 47.64 |  |
|  | Median | 48.84 |  |
|  | Variance | 236.24 |  |
|  | Std. Deviation | 15.37 |  |
|  | Minimum | 19 |  |
|  | Maximum | 73 |  |
|  | Range | 54 |  |
|  | Interquartile Range | 21 |  |
|  | Skewness | -0.162 | 0.374 |
|  | Kurtosis | -0.7 | 0.733 |

Breast cancer is the most common among female cancers, and both its baseline prevalence and incidence exceed that of other types of cancer in both genders combined, Table 2. The second most common type of cancer was thyroid cancer, which had 24 cases in females and 7 in males, with a total of 31 cases. The third was colorectal cancer, with a total of 24 cases, which was 10 and 14 in both females and males, respectively. The fourth most common type of cancer was the prostate, which ranked the top cancer in males with 22 cases. Other types of cancer had a variable number of cases among genders, ranging from 13/12 cases in liver and cervical and only to 1 case in gallbladder and peritoneum.

Table 3 compares this study’s results with the latest published UAE cancer national registry in 2019. Starting with breast cancer, the percentage of cases in our data was 30% in females, with a close rate of 28% in the registry. A similar finding was observed in thyroid cancers, with 18.6% and 16.3% in females and 6.3% and 5.5% in males. There was a minimal difference of 0.9% in colorectal cancer in females and 1.5 in males. Other significant similarities were found in the rest of the types of cancer, as in Table 3. All confirm the resemblance between this study and the latest national registry.

With regard to the age distribution of all types of cancer among females, Figure 1 shows that the peak prevalence of cancer among females is between the ages of 40 and 60, and the median for female participants with a negative history of cancer after WEQAYA was 36.5. These numbers reflect the young population of the UAE. The median age with a positive history was 62.1. In males, it shows a skewed left with the highest peak between 60 and 80 years. The male median for participants with a negative history of cancer was 35.4, and for those with a positive history, it was 47.52. Table 4 shows the median age of history of cancer at the time of screening; the median for female participants with a negative history of cancer was 36.8, while the median age with a positive history was 62.7. Among males, the median for participants with a negative history of cancer was 35.7 years, while the median age with a positive history was 48.84.

**Figure 1.**
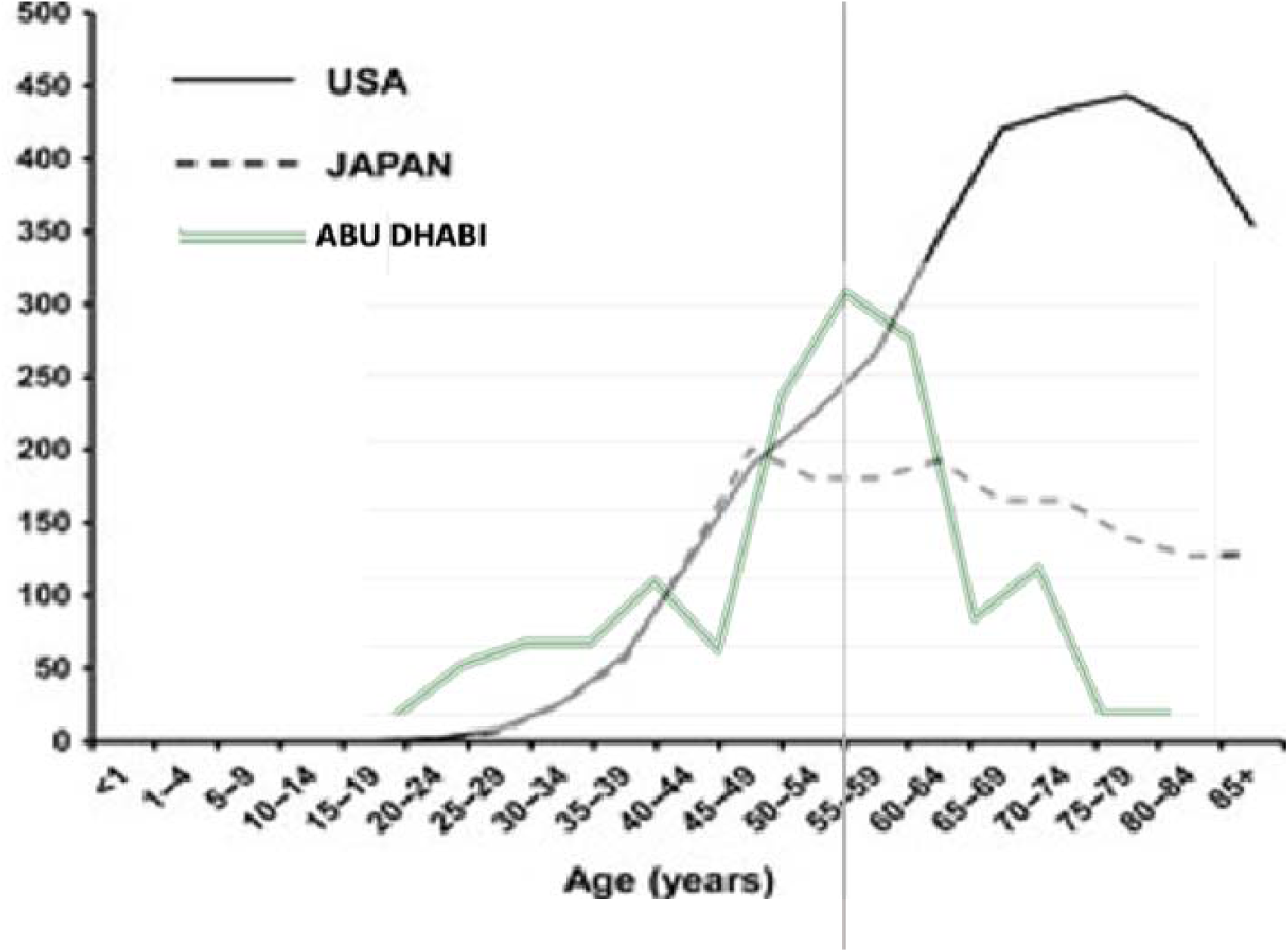
Prevalence of cancer in Abu Dhabi in this cohort compared to the US and Japan distributed by age

**Figure 2.**
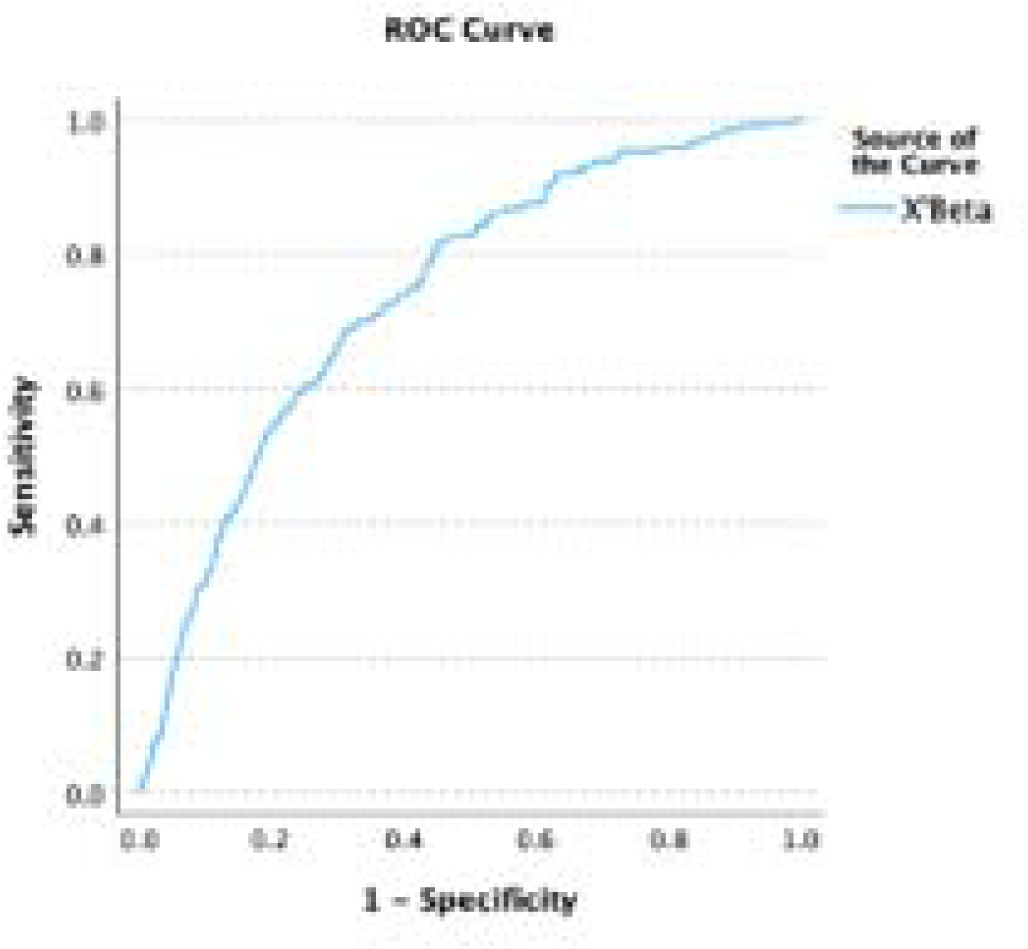

Among the whole cohort, Cox regression analysis showed only age and higher levels of HDL as risk factors for any cancer after screening when including possible risk factors that were assessed at baseline. The risk increases by 4.8% for each year of age and 52% for each unit increase in HDL. Figure 2 The area Under the ROC Curve for predicting any cancer type is 0.739 (0.707-0.771). Figure 3

## Discussion

This study shows a great similarity between our findings and those of the national registry, supporting the validity of both data sources. With regard to changing trends in cancer epidemiology, a study done in the UAE, Al Ain, in 1997[5] showed that the most common male malignancy was gastric carcinoma (17.6%), followed by lymphoma (16.8%), colorectal cancer (14.9%) and skin cancer (8.3%). In contrast, in this study, the most common male malignancy was prostate (20%), followed by colorectal (12.7%), liver (9%), and thyroid (6.3%) and lymphoma (6.3%).

In comparison, the most common cancer in females in the 1997 study was breast (31.6%), followed by cervix (27%), thyroid (8.3%), and colorectum (7.9%). In this study, breast cancer was also the most common in the female group. However, the second most common was thyroid, followed by cervical and colorectum. This change in prevalence in males could reflect the increase in some cancer types, such as colorectal carcinoma. A more detailed study of the risk factors of individual cancer types is essential to inform us of any preventive measures.

In the systematic analysis for the Global Burden of Disease Study, metabolic risk factors of cancer saw the largest increases between 2010 and 2019 [2]. While there is mounting evidence that increasing the level of HDL-C can reduce cardiovascular risk [6], in this study, it is associated with an increased risk of all cancer types. This relationship is not well understood, and variations in results, depending on the type of cancer, were observed. In a large population-based cohort in Korea, Low HDL-C increased the risk of gastric cancer, whereas high HDL-C was associated with esophageal cancer risk with discrepancies by sex and smoking status [7,8]. In contrast, low and highly high HDL-C levels compared to normal levels were associated with an increased liver cancer risk. The unfavorable association between low HDL-C and liver cancer was remarkable in smokers and pre-menopausal women [9]. Although a hallmark of cancers is altered lipid metabolism, changes in lipid metabolism have received less attention than other changes in cancer cells. Recent studies have demonstrated a relationship between lipid reprogramming and cancer progression.

Genetic bases for this are also suggested, with the SCARB1 gene also encoding the scavenger receptor class B type I (SR-BI), which is important in regulating cholesterol exchange between cells and high-density lipoproteins. Solid tumors have been shown to accumulate more cholesterol than the host healthy tissues. Although no study examining the role for SR-BI in the promotion of cell transformation has been performed, it has been recognized that cancer cells can use the HDL/SR-BI pathway to take up cholesteryl ester and enhance malignant phenotypes. The role of HDL/SR-BI in breast and prostate cancers has been the most extensively studied, and over-expression of SR-BI can enhance HDL-mediated proliferation of the breast cancer cell, and down-regulation of SR-BI has been shown to cause a significant reduction in cellular viability [10, 11].

This suggests that lipid metabolism has high potential as a novel biomarker for the diagnosis, prognosis, and therapy of cancer [12]. It has been suggested that [13] manipulating their levels might provide novel, efficient therapeutic strategies for treatment [14].

Statin use was associated with decreased CRC risk, and high medication compliance was inversely associated with CRC risk in patients with and without dyslipidemia compared to non-use of medication. The UAE is among the best countries in the world to investigate the impact of statins on cancer risk due to the high level of statin use, and both conditions are very prevalent in the UAE population [15]. This is another area where cancer risk may be modified. Regular health examinations can help identify individuals who are vulnerable to CRC, and continued statin use may lower the risk of CRC [16].

Later studies identified hyperinsulinemia, elevated IGFs, hyperglycemia, dyslipidemia, adipokines, inflammatory cytokines, and the gut microbiome [17] as potentially modifiable risk factors. Modifying such risk factors, such as intentional weight loss, may protect against cancer development, and therapies for diabetes may prove to be effective adjuvant agents in reducing cancer progression [18,1].

This study’s limitation is the relatively modest sample size of 239 individuals with a history of cancer. Another limitation is that other important risk factors, such as positive family history, reproductive history, and hormone replacement history, are unavailable. Its major strength is that it is a large community-based cohort study with a comprehensive review of the reported prevalence of a history of cancer among adult UAE nationals in Abu Dhabi/Al Ain over a mean follow-up of 9.4 years. This study underscores the importance of studying more risk factors with more interventions to investigate the uniqueness of cancer risk factor interaction in this population. This will enable better development and selection of diagnostic and therapeutic options. Finally, developing risk estimation is also critical for cancer prevention and early detection, and this study is the first in the UAE and the region.

## Conclusion

Increased HDL levels and older age were the identified risk factors when all cancer types were studied together. This study calls for further research on different types of cancers to identify significant predictors for possible early diagnosis and management. It also calls for research targeting risk factor modification and its effect on lowering cancer prevalence and survival. This study’s results are critical to informing the country’s decision-making in planning cancer services. They are informative internationally as the risk factors identified are prevalent worldwide and can be targeted with available therapeutics.

## Data Availability

All data produced in the present study are available upon reasonable request to the authors

## Declarations

### Ethical approval and consent to participate

The study was approved by the AlAin Human Ethics Committee, approval number 13/58, and Ambulatory Healthcare Services IRB 19-2022. All methods were carried out under relevant guidelines and regulations. The authors confirm that the study was conducted in accordance with the Helsinki Declaration.

### Consent statement in the Ethics approval and consent to participate

Informed consent was waived by the IRBs as the study was designed for retrospective data gathered as part of patient care and anonymized at analysis.

### Authors’ contributions

LBK and NN conceptualized and analyzed data. LBK, RA, and MAA wrote the manuscript; all other co-authors collected data and reviewed the manuscript. All authors have read and approved the final manuscript.

### Consent to publish

Not Applicable.

### Availability of data and materials

Data availability is restricted due to institution policies.

## Acknowledgments

None.

